# DermNet: An Incremental Knowledge Distillation Framework for Accurate and Robust Skin Layer Segmentation

**DOI:** 10.64898/2026.11.11.26367811

**Authors:** Anum Abdul Salam, Muhammad Usman Akram, Soyiba Jawed, Tehmina Shehryar, Zoha Zahid Fazal, Muhammad Haroon Yousaf, Babar Rao, Norah Saleh Alghamdi

## Abstract

**Background:** Accurate segmentation of skin layers in histopathological whole-slide images (WSIs) is important for identifying disease-associated structural changes and supporting computer-aided dermatopathological analysis. However, existing approaches are often restricted to single magnification levels and may inadequately capture the heterogeneous morphological characteristics of individual skin layers. We propose DermNet, a multi-resolution class-incremental learning framework designed to achieve accurate skin layer segmentation while mitigating catastrophic forgetting.

**Methods:** DermNet employs a SegFormer backbone within a teacher–student architecture and progressively learns class-specific features from WSIs acquired at 10×, 5×, and 2× magnifications. The framework incorporates exemplar replay, mutual knowledge distillation, classification and Kullback–Leibler divergence losses, boundary loss, and Elastic Weight Consolidation to preserve previously learned representations while incorporating new tissue classes. The model was trained and evaluated using a publicly available dataset of 290 WSIs, with segmentation focused on the epidermis, papillary dermis, reticular dermis, hypodermis, keratin, and background.

**Results:** DermNet consistently outperformed U-Net and alternative SegFormer training strategies across all three magnification levels. At 10× magnification, DermNet achieved Dice scores of 0.95 for epidermis, 0.92 for papillary dermis, 0.94 for reticular dermis, 0.97 for keratin, and 0.98 for hypodermis, outperforming UNet++, DeepLabv3+, TransUNet, SwinUNet, and nnU-Net under the same experimental conditions. Evaluation on an independent publicly available dataset further demonstrated cross-dataset generalizability, with Dice scores ranging from 0.89 to 0.97 across classes.

**Conclusion:** DermNet enables accurate and robust multi-resolution skin layer segmentation by combining class-incremental learning with knowledge distillation and mechanisms for preserving previously learned features. Its performance across magnification levels and an independent dataset supports its potential for scalable and generalizable automated dermatopathological image analysis.

## Introduction

Dermatological diseases range from benign inflammatory conditions to aggressive malignancies and represent a substantial clinical burden. Histopathological examination of hematoxylin and eosin (H&E)-stained skin biopsies remains central to definitive diagnosis; however, interpretation is time-consuming, requires specialized expertise, and may be affected by interobserver variability^1,2^. Advances in whole slide imaging (WSI), computer-aided diagnosis (CAD), and deep learning have therefore created opportunities to automate and support dermatopathological assessment^3–5^.

Accurate segmentation of skin layers is particularly important because the epidermis, dermis, and hypodermis exhibit distinct structural and pathological alterations. Identification of these tissue compartments can facilitate localization and characterization of disease-associated features and provide clinically meaningful information for automated histopathological analysis^6,7^. Semantic segmentation of clinically relevant structures in skin biopsy WSIs has consequently emerged as an important component of computational dermatopathology^7,8^. **Figure 1** illustrates the principal anatomical layers of the skin alongside their corresponding appearance in H&E-stained tissue.

**Figure 1.**
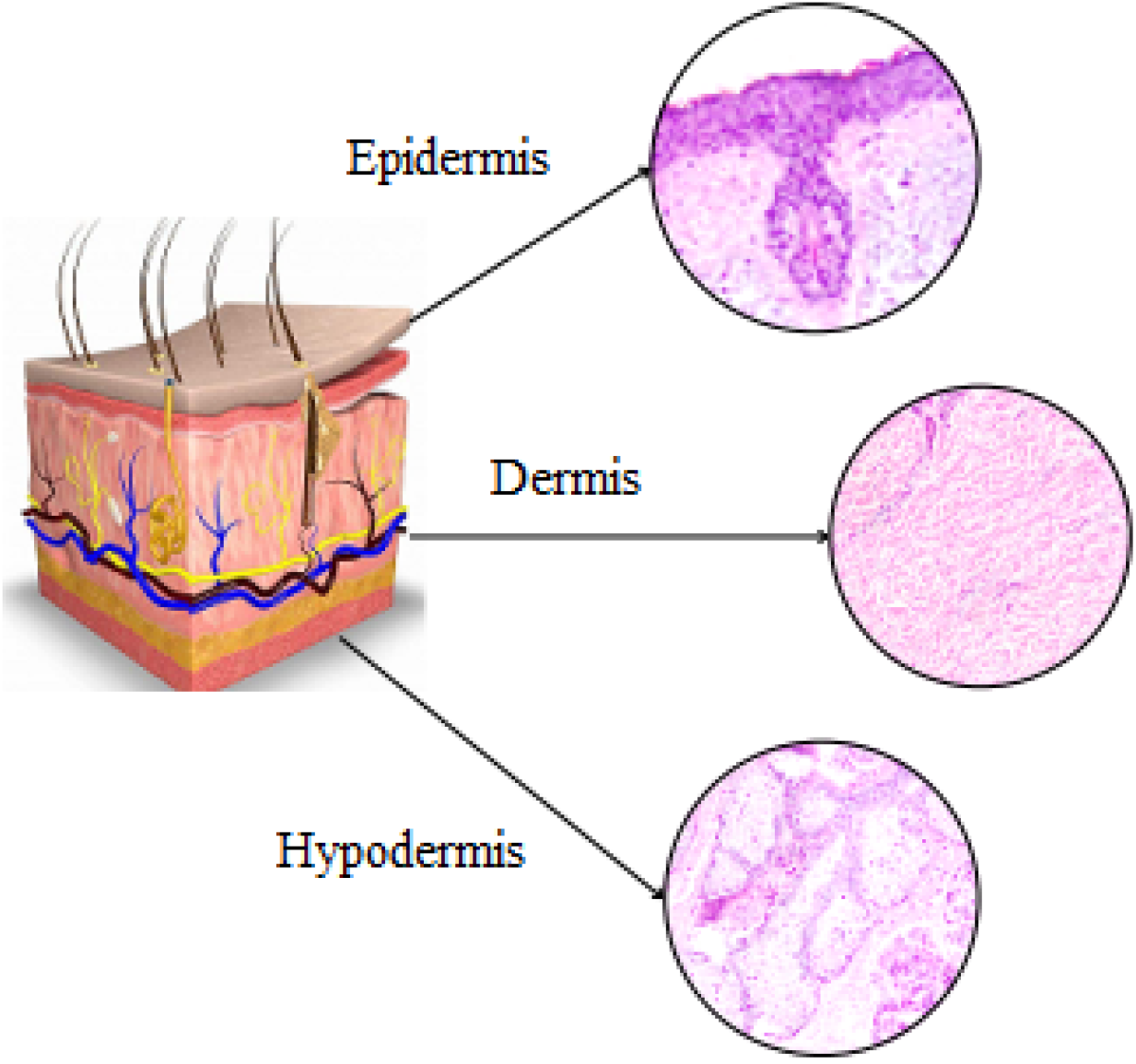
Anatomical and histological representation of the principal skin layers. Schematic cross-section of human skin illustrating the epidermis, dermis, and hypodermis alongside representative H&E-stained histological regions, highlighting the distinct tissue morphology of each layer.

Brightfield microscopy enables pathologists to examine histological specimens at different magnifications, integrating low-magnification tissue architecture with higher-magnification morphological detail. Similarly, digital WSI provides multiresolution representations of histological slides that can support both computational analysis and diagnostic interpretation^3,9^. However, automated analysis of WSIs remains challenging because of their large dimensions, heterogeneous tissue morphology, and the substantial effort required to generate detailed pixel-level annotations^7,10^.

Deep learning has demonstrated considerable potential for segmentation and classification of dermatopathology WSIs, including basal cell carcinoma (BCC), squamous cell carcinoma (SCC), melanoma, and clinically relevant skin structures^8,11^. Nevertheless, existing approaches frequently focus on specific lesions, tissue structures, or individual imaging scales, leaving an opportunity for frameworks that can progressively learn multiple skin layers while retaining previously acquired information and operating across different magnifications. Therefore, we propose DermNet, a class-incremental deep learning framework for segmentation of skin layers from WSIs acquired at 10×, 5×, and 2× magnifications, designed to support multi-resolution learning while mitigating catastrophic forgetting.

## Materials and Methods

### Dataset

The dataset utilized in this study is a publicly available repository comprising 290 WSI samples, including 100 shave biopsies, 58 punch biopsies, and 132 excisional biopsies. The histological slides were obtained from patients aged 34–96 years, with a median age of 70 years. Females constituted approximately two-thirds of the cohort, while males comprised approximately one-third.

Images were captured using a brightfield microscope at three magnification levels: 10×, 5×, and 2×. Each sample was associated with a ground-truth image labeled for 12 classes, including background (BKG); skin layers comprising epidermis (EPI), hypodermis (HYP), papillary dermis (PAP), and reticular dermis (RET); other tissue and structural features including follicle (FOL), keratin (KER), and inflammation (INF); and carcinomas comprising BCC, intraepidermal carcinoma (IEC), and SCC. Representative H&E-stained WSIs at the three magnification levels, together with their corresponding ground-truth segmentation masks, are presented in **Figure 2**.

**Figure 2.**
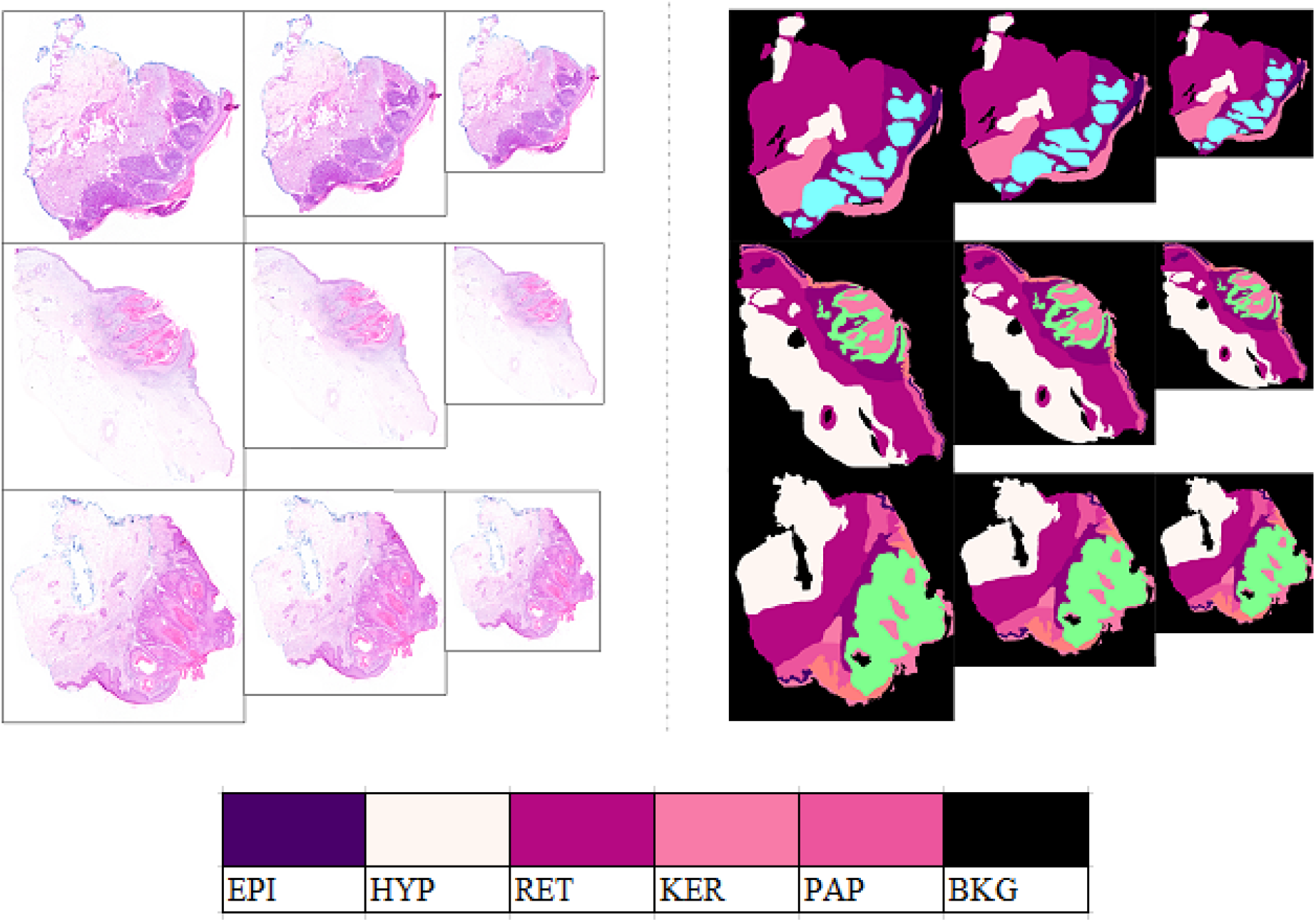
Representative samples from the Queensland dataset at multiple magnification levels with corresponding ground-truth segmentation masks. The left panel shows H&E-stained whole-slide images at 10×, 5×, and 2× magnifications, while the right panel shows the corresponding annotated segmentation masks. Colors represent epidermis (EPI), hypodermis (HYP), reticular dermis (RET), keratin (KER), papillary dermis (PAP), and background (BKG).

For the present study, the ground-truth annotations were further modified to retain only the classes relevant to skin layer segmentation: epidermis, papillary dermis, reticular dermis, hypodermis, and keratin, while the remaining regions were assigned to the background class. The representation ratio of each retained class in the utilized dataset is presented in **Table 1**.

**Table 1.**
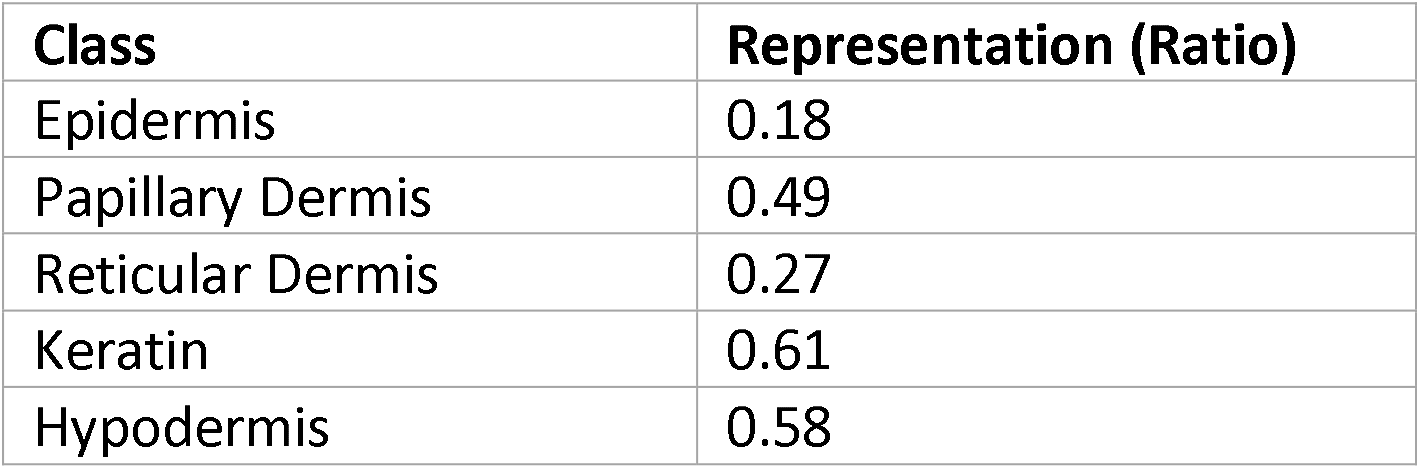
Representation ratio for each class.

| **Class** | **Representation (Ratio)** |
|---|---|
| Epidermis | 0.18 |
| Papillary Dermis | 0.49 |
| Reticular Dermis | 0.27 |
| Keratin | 0.61 |
| Hypodermis | 0.58 |

In incremental learning, to avoid catastrophic forgetting, it is essential to replay some exemplar instances of classes learned in the previous stage *S*_*i−* 1_ to be replayed in the current stage *S*_*i*_ . As illustrated in **Figure 3**, for the first stage we trained the model on 50% of instances of epidermis from the dataset. In the second stage, to train the student model, we utilized 50% samples of the hypodermis (papillary dermis, reticular dermis) along with 25% instances of epidermis to prevent catastrophic forgetting. Finally, in the third stage, 100% samples of keratin and hypodermis, along with 50% samples of papillary and reticular dermis and 25% samples of epidermis were utilized in model training. This exemplar replay strategy reinforces previously learned class representations while progressively incorporating new classes, thereby mitigating catastrophic forgetting. Data from all three magnification levels (10×, 5×, and 2×) were incorporated across the incremental training stages to facilitate multi-resolution feature learning from coarse to fine scales.

**Figure 3.**
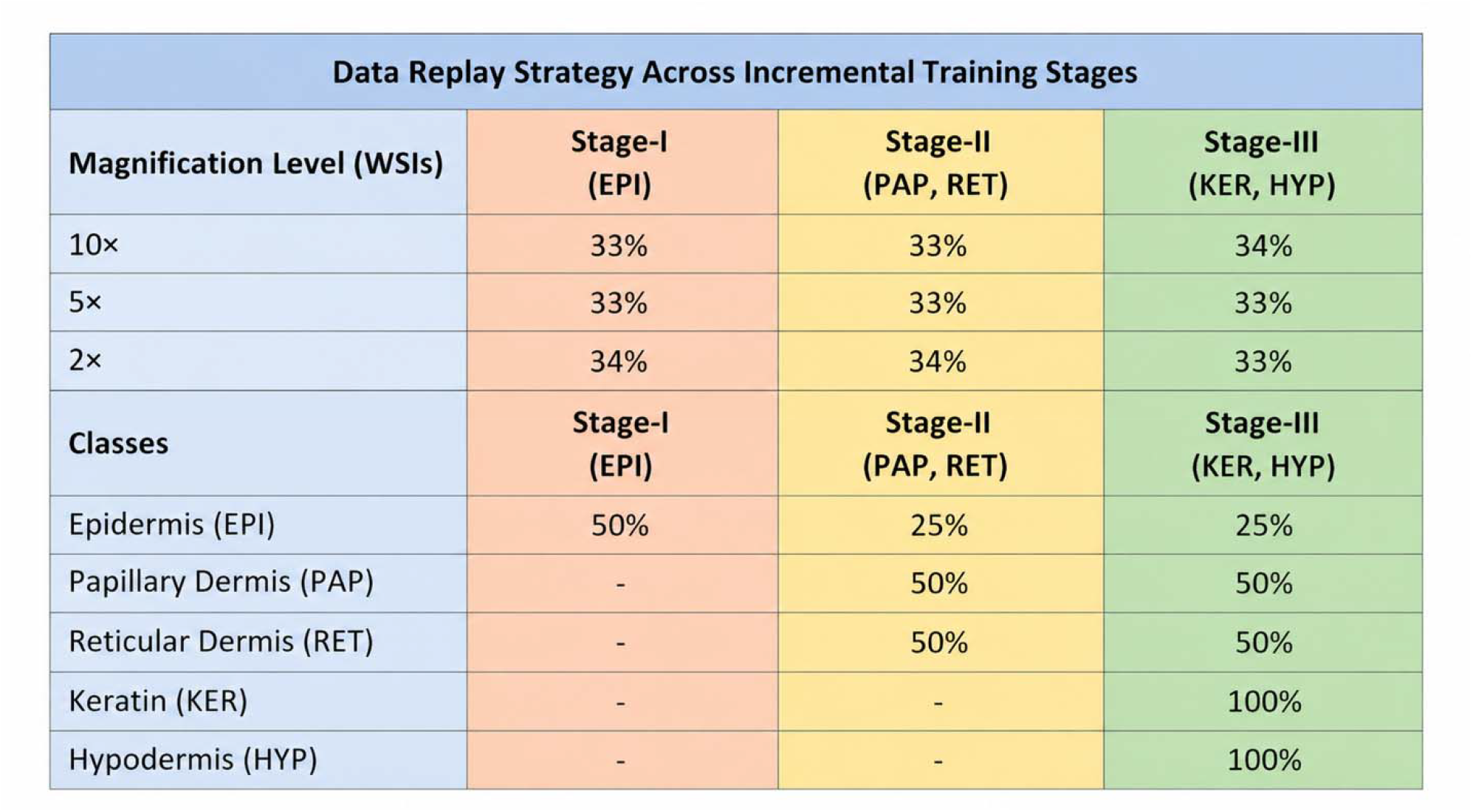
Data replay strategy across the three stages of DermNet incremental training. Whole-slide image (WSI) samples acquired at 10×, 5×, and 2× magnifications were incorporated across all training stages, with approximately equal representation from each magnification level. Class-specific exemplar replay was performed progressively: Stage I included 50% of epidermis (EPI) samples; Stage II included 25% EPI and 50% each of papillary dermis (PAP) and reticular dermis (RET) samples; and Stage III included 25% EPI, 50% each of PAP and RET, and 100% each of keratin (KER) and hypodermis (HYP) samples. This progressive replay strategy was designed to facilitate acquisition of new class-specific features while preserving previously learned representations.

To address the challenge of the high-resolution dataset, where each image is comprised of approximately 100,000 pixels, we have adopted a patch-based approach. Before model training, the dataset was pre-processed into non-overlapping 256×256 patches. This method has helped us reduce memory consumption and computation time. Furthermore, the patch-based technique has also allowed the model to focus on specific regions, allowing it to gather contextual features more precisely thereby improving overall segmentation performance.

Exemplar data replay approach for incremental learning with three steps. To enable multi-resolution learning, the dataset is replayed at varying magnification levels (10×, 5×, and 2×). Furthermore, the training focus gradually moves across various skin layers (epidermis, papillary dermis, reticular dermis, keratin, and hypodermis), providing progressive learning and accurate segmentation.

### Methodology

Instead of training the entire model in a single stage, we propose a multi-staged training process via a student-teacher model. This technique enables concentrated and systematic learning of the various skin layers marked in the ground truth, allowing the model to adequately represent the intricacies of each layer. Note that the dataset consists of WSIs at different magnification levels and hence during training, all magnifications should also be incorporated into the model. This multi-resolution technique allows the model to use both the wider contextual information from lower magnifications and the finer details from higher magnifications, resulting in thorough and accurate segmentation across all skin layers.

Figure 4. illustrates the overall DermNet framework, including the three-stage class-incremental teacher–student training strategy and the subsequent inference pipeline for automated skin layer segmentation from WSIs.

### Classwise Incremental Learning

In our conducted research we have divided our training phase into three stages where each stage depicts class incremental learning. In the training stage, 75% images contain papillary dermis and reticular dermis, and the remaining 25% images contains epidermis samples to ensure the model retains the features of previously learned classes. Finally, keratin and hypodermis have larger regions with more distinct features, making them suitable for learning in later stages without negatively affecting retention of more intricate classes. Figure 5 illustrates the complete training pipeline, clearly highlighting the training data passed and weight regularization at each training stage. As elaborated in the figure, at each training stage model has been trained for targeted classes using images of various magnification levels, thus making the trained model more robust towards change in magnification levels.

**Figure 4.**
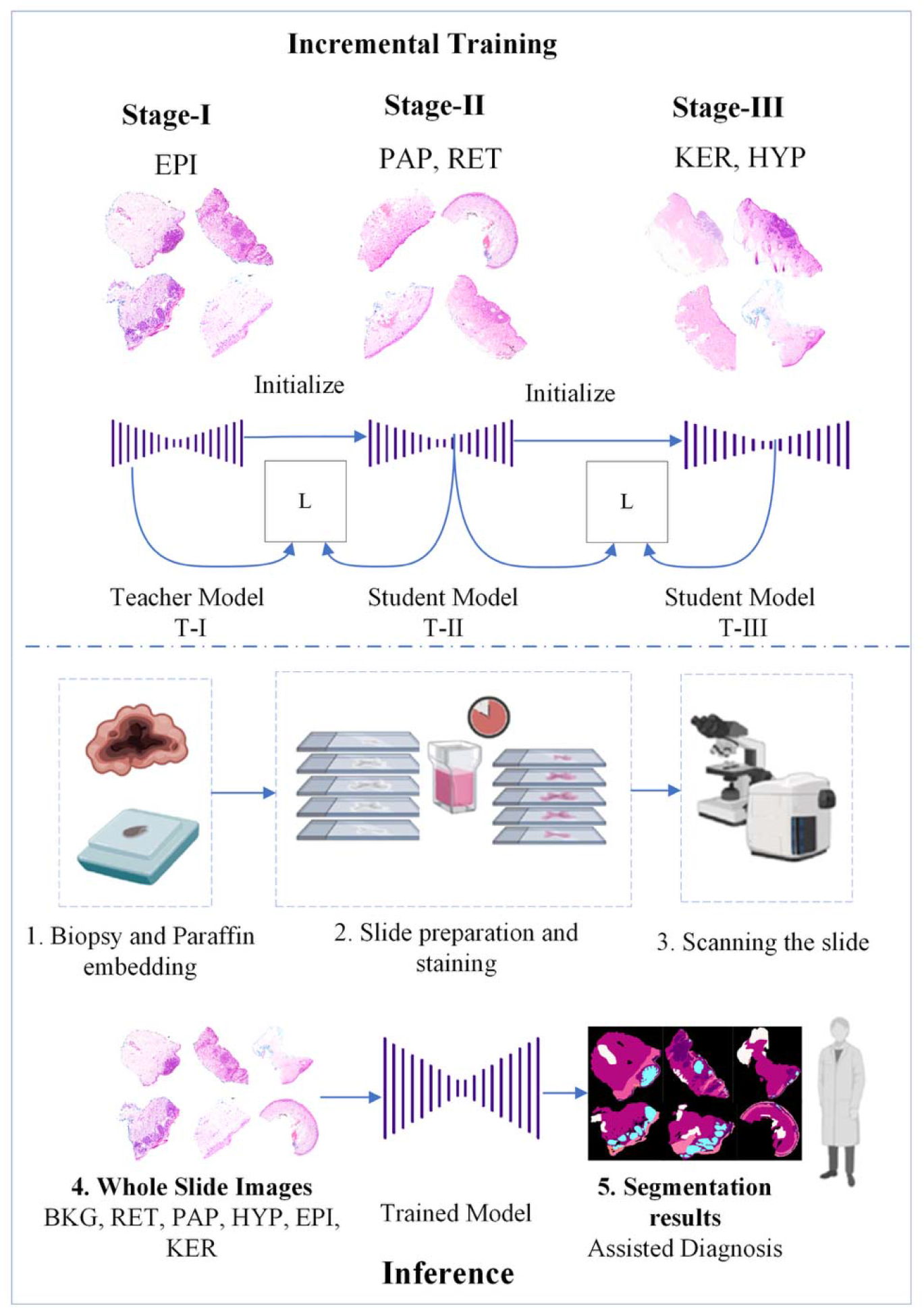
Overview of the proposed DermNet framework for incremental skin layer segmentation and inference. The upper panel illustrates the three-stage class-incremental training strategy, in which the model progressively learns epidermis (EPI), papillary dermis (PAP) and reticular dermis (RET), followed by keratin (KER) and hypodermis (HYP), using a teacher–student architecture and knowledge transfer between successive stages. The lower panel illustrates the inference workflow, from biopsy preparation, staining, and whole-slide image acquisition to processing by the trained model and generation of class-specific segmentation maps for assisted diagnosis.

**Figure 5.**
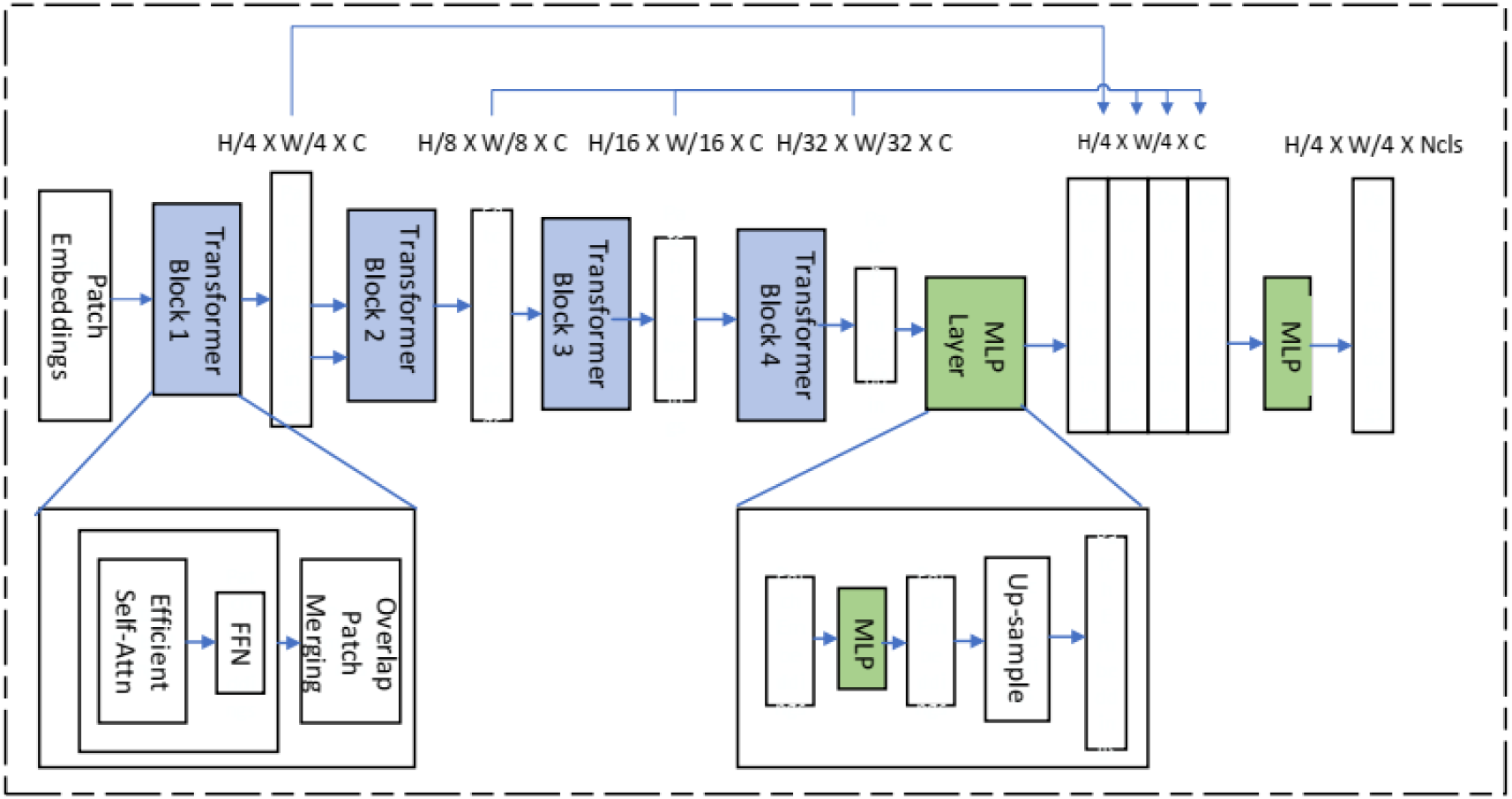
Architecture of the SegFormer backbone employed in the DermNet framework. The hierarchical Transformer encoder progressively extracts multi-scale feature representations through overlapping patch embeddings and Transformer blocks, while the multilayer perceptron (MLP)-based decoder integrates features from different resolutions to generate the final semantic segmentation output.

### Model Training and weight regularization

To train the teacher model in stage-I we have utilized a SegFormer backbone pre-trained on ADE20K dataset. The architecture of the SegFormer backbone employed in DermNet is illustrated in **Figure 5**, highlighting its hierarchical Transformer encoder and multi-scale feature aggregation for semantic segmentation. Once the teacher model was trained, the student models were optimized using a combination of mutual distillation loss, classification loss, Kullback–Leibler (KL) divergence loss, and boundary loss to facilitate incremental learning while mitigating catastrophic forgetting.

We employed cross-entropy loss to optimize model convergence, as represented in **Equation 1**.

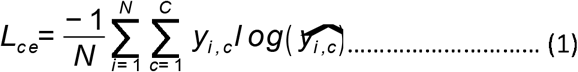

Where *N* indicates the number of images, *C* annotates the total number of classes, *y*_*i, c*_ indicates the ground truth of *c*^*t h*^ class for *i*^*th*^ image sample, and *ŷ* _*i, c*_ indicates the predicted probability. Once the teacher model has been trained, to ensure learning without forgetting we have trained the student model using a combination of mutual distillation loss *L*_*md*_, classification loss *L*_*c*_, KL *L*_*K L*_ loss and boundary loss *L*_*bound ar y*_ . **Equation 2** highlights the combined loss function used to converge student models via incremental learning.

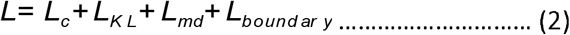

Classification loss *L*_*c*_ incorporates cross-entropy loss to ensure the model learns to segment all the classes present in ground truth at each stage. It enforces the model to learn new classes while remembering the previously learned classes at each stage. *L*_*c*_ is represented as **Equation 3**.

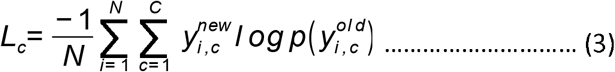

Where *y*^*new*^ indicates the sample from the new class and *y*^*ol d*^ indicates the sample from the class learned at the previous stage. To identify the structural and semantic similarities between the newly learned classes and previously learned classes, mutual distillation loss *L*_*md*_ has been employed which works on Bayesian inference, regularizing samples from both newly learned classes and previously learned classes as evident in **Equation 4**.

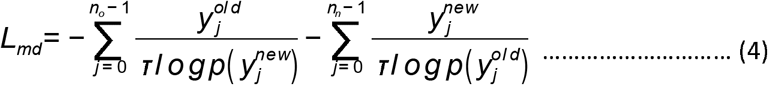

Where τ indicates the scaling factor and *n*_*o*_ and *n*_*n*_ represents the number of old and new samples, respectively. To further enhance the prediction accuracy, we have implemented KL loss *L*_*K L*_ to compare and reduce the difference between ground truth and predicted samples at each stage. *L*_*K L*_ can be mathematically represented by **Equation 5** as:

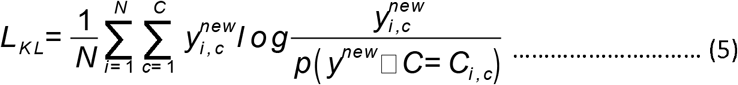

The boundary loss aims to improve the segmentation performance at the layer boundaries, which is essential for precise skin layer segmentation in WSIs. The transitions between these thin and anatomically significant layers are precisely localized with the help of boundary loss, which prevents them from being confused or incorrectly classified. Mathematically, it can be represented by **Equation 6** as:

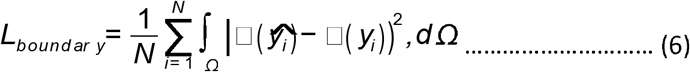

Here,^Ω^ indicates the image’s spatial domain, and □( *y*_*i*_ ) and □ (*ŷ* _*i*_ ) stands for the gradients of the ground truth and predicted segmentation masks, respectively. To guarantee alignment, the loss reduces the discrepancy between the ground truth and predicted boundaries. Moreover, to regularize the weights before updating them in each training stage, we have employed Elastic Weight Consolidation (EWC) using the Fisher Information Matrix (FIM) to avoid catastrophic forgetting. EWC consolidates the weights significant for the previous tasks while updating the rest of the weights by employing a regularization term in the loss function mathematically represented in **Equation 7**.

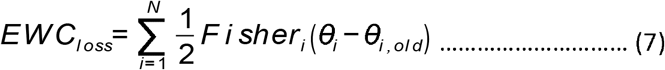

where θ_i_ and θ_*i,old*_ are the weight matrices at *i*^*th*^ index for the current and previous task. *Fi sher* _*i*_ holds the importance of weight θ_*i*_ for previous tasks computed using **Equation 8**.

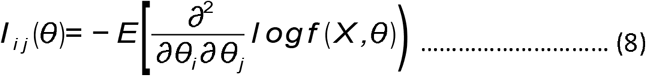

The Fisher information matrix (FIM) is calculated using the second derivative of log-likelihood as represented in **Equation 9**, Where *f* ( *X*,θ) indicates the probability density function of random variable *X, E*[.) denotes the expectation operator.

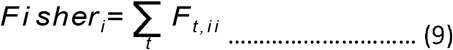

Where *F*_*t,i i*_ is the (*i,i* ) element of the Fisher information matrix ( *I* ) for *t* tasks, where each element (*i, j* ) of the matrix is calculated using **Equation 7**.

### Post Processing

In the post-processing stage of skin layer segmentation, we perform several actions to improve the output. The method begins with connected component labeling, in which each linked region in the binary segmentation map is labeled with eight connected neighbors. Size thresholding removes noise by identifying and merging regions with pixel counts below a threshold (*T*_*mi n*_), where *T*_*mi n*_ is set to 50 pixels based on experimental evaluation to remove small, isolated regions without affecting meaningful structures. The hole-filling operation fills in minor gaps in the segmentation results utilizing morphological operations such as dilation, represented by:

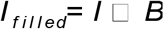

Here *I* represents the binary image, *B* is the structuring element (a square kernel), and *x*□*y* represents the dilation operation. The size of the structuring element *B* is set to 3*×* 3, as this provides a balance between filling gaps and avoiding excessive merging of regions. Following hole filling, the result is improved using a Gaussian smoothing filter to decrease noise and smooth borders. This may be mathematically phrased as:

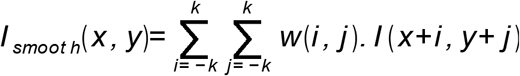

where *w* (*i, j* ) is the weight of the Gaussian kernel, defined as:

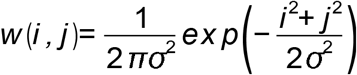

σ determines the kernel’s spread. The size of the Gaussian kernel is chosen as *k*= 3 (resulting in a 7*×* 7 kernel), and ^σ^ is set to 1.0. These values ensure sufficient smoothing of the segmentation map while retaining important boundary details. These post-processing steps help to enhance the segmentation results, resulting in more accurate segmentation of skin layers and fewer artifacts in the final segmentation map.

### Experimentation and Results

### Experimental Setup

The proposed framework was implemented using **Python 3.11.4 and PyTorch** and trained on an NVIDIA GeForce RTX 2070 GPU. For Stage I, the teacher model employed a SegFormer backbone pretrained on the ADE20K dataset. The models were trained using the Adam optimizer with a batch size of 9 for 6 *X* 10^*−* 5^ learning rate. The teacher model was trained for 100 epochs, whereas the student models were trained for 50 epochs at each subsequent incremental stage. Data augmentation involving variations in image scale and orientation was applied to reduce overfitting and improve robustness. The dataset was divided into 70% training and 30% testing subsets.

### Evaluation Metrics

Segmentation performance was evaluated using accuracy, mean Intersection over Union (mIoU), F1-score, precision, Hausdorff Distance (HD), and Boundary Dice (BD-Dice). Accuracy, mIoU, F1-score, and precision quantify pixel- and class-level segmentation performance, whereas HD and BD-Dice provide complementary assessments of boundary agreement between predicted and ground-truth segmentation masks. The assessment metrics listed above are mathematically expressed as follows:

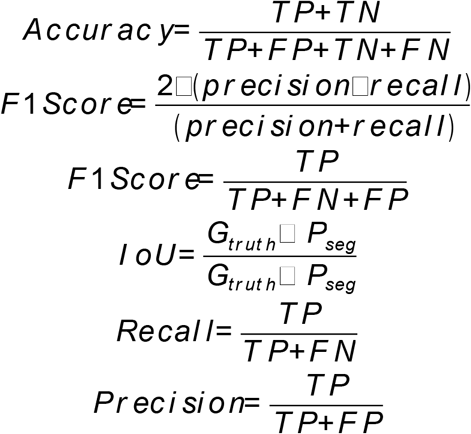

$$\begin{aligned} HD(X,Y) &= \max\{d_{XY}, d_{YX}\} \notag \\ &= \max\left\{\max_{x \in X} \min_{y \in Y} d(x, y), \max_{y \in Y} \min_{x \in X} d(x, y)\right\} \end{aligned}$$

Where *T P, F P, T N*, and *F N* are True positives, False Positives, True Negatives, and False Negatives respectively. Ground truth is indicated as *G*_*tr ut h*_, and prediction from the trained model is represented as *P*_*seg*_. *d* annotates the euclidean distance computed between ground truth pixels *x* and segmentation prediction *y* .

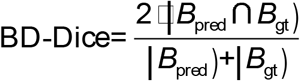

Here, *B*_pred_ and *B*_gt_ denote the boundary pixel sets of the predicted and ground-truth masks, respectively. BD-Dice captures fine-grained discrepancies at region borders.

Because DermNet employs class-incremental learning, forward transfer (FWT), backward transfer (BWT), and forgetting score (FS) were additionally evaluated. FWT measures the extent to which previously acquired knowledge facilitates performance on subsequently introduced tasks, whereas BWT quantifies the influence of learning new tasks on previously learned tasks. Negative BWT indicates deterioration in performance on earlier tasks. FS quantifies the maximum performance decrease for a previously learned task following the introduction of subsequent tasks. Together, these metrics provide complementary measures of knowledge transfer and retention throughout incremental training^12^ .

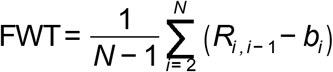

where *R*_*i, i−* 1_ is the accuracy on task *i* immediately after training on task *i −* 1, and *b*_*i*_ is the initial performance on task *i* before training.

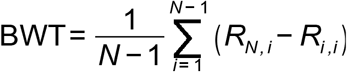

where, *R*_*N, i*_ is the final performance on task *i* after all tasks have been learned, and *R*_*i, i*_ is the performance on task *i* immediately after it was learned.

FS measures the maximum performance drop experienced by a task due to learning subsequent tasks.

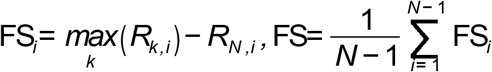

These metrics provide a comprehensive understanding of how well the model retains and transfers knowledge across stages during class-incremental learning.

### Ablation Study

To investigate the contribution of the proposed incremental-learning strategy, three training configurations were evaluated. Model-I consisted of a SegFormer model trained using conventional transfer learning and fine-tuning. Model-II (FL) incorporated incremental learning with selective freezing and fine-tuning of the SegFormer backbone. Model-III (DermNet) represented the complete proposed incremental-learning framework.

For Model-II (FL), layer freezing was progressively applied to balance retention of previously learned features with adaptation to newly introduced classes. During Stage I, the complete SegFormer encoder and decoder were trained on the initial dataset. During Stage II, the early encoder layers were frozen to retain lower-level representations, while later encoder layers responsible for higher-level semantic information were fine-tuned and the decoder remained fully trainable. During Stage III, both early and intermediate encoder layers were frozen, whereas the later encoder layers and decoder continued to be trained.

In contrast, DermNet retained all model layers as trainable throughout the incremental stages, while the proposed distillation and regularization strategy was used to preserve previously acquired information. This design allowed both lower- and higher-level feature representations to continue adapting as new tissue classes were introduced.

### Comparison with the Baseline Model

The first set of experiments compared DermNet with a U-Net-based skin segmentation model, the conventionally fine-tuned SegFormer (Model-I), and the incrementally trained SegFormer with selective layer freezing (Model-II [FL]). Class-wise segmentation performance was evaluated at 10×, 5×, and 2× magnifications.

As presented in **Table 2**, DermNet achieved the strongest overall class-wise segmentation performance across the three magnification levels. Model-I showed comparatively lower performance for classes with smaller pixel representation, particularly EPI and PAP. Model-II (FL) improved upon Model-I across the evaluated classes and magnifications, indicating better preservation of previously acquired representations during sequential training. However, DermNet further improved segmentation of challenging tissue classes, including EPI, PAP, keratin (KER), and reticular dermis (RET), while maintaining performance across different magnification levels.

**Table 2.** Class-wise Accuracy comparison of the proposed framework (DermNet) with U-Net model trained and tested on Queensland dataset, baseline model (Model-I), and incrementally learned model with frozen layers in stage II and stage III.

| Methodology | Dataset | BKG | EPI | PAP | RET | KER | HYP |
|---|---|---|---|---|---|---|---|
| UNet |  | 0.95 | 0.83 | 0.80 | 0.70 | 0.84 | 0.96 |
| Model-I |  | 0.93 | 0.65 | 0.68 | 0.67 | 0.80 | 0.90 |
| Model-II(FL) | 10x | **0.98** | 0.89 | 0.85 | 0.87 | 0.81 | 0.96 |
| DermNet |  | **0.98** | **0.95** | **0.92** | **0.94** | **0.97** | **0.98** |
| UNet |  | 0.93 | 0.84 | 0.78 | 0.81 | 0.81 | 0.95 |
| Model-I |  | 0.93 | 0.67 | 0.70 | 0.71 | 0.85 | 0.88 |
| Model-II(FL) | 5x | **0.98** | 0.88 | 0.87 | 0.87 | 0.92 | 0.94 |
| DermNet |  | **0.98** | **0.91** | **0.90** | **0.95** | **0.96** | **0.92** |
| UNet |  | 0.95 | 0.74 | 0.77 | 0.68 | 0.87 | 0.96 |
| Model-I |  | 0.93 | 0.70 | 0.71 | 0.73 | 0.88 | 0.90 |
| Model-II(FL) | 2x | **0.98** | 0.86 | 0.89 | 0.85 | 0.90 | 0.91 |
| DermNet |  | **0.98** | **0.90** | **0.92** | **0.91** | **0.94** | **0.90** |

DermNet was subsequently compared with several established segmentation architectures, including UNet++, DeepLabv3+, TransUNet, SwinUNet, and nnU-Net, with all models trained and evaluated under the same experimental conditions on the Queensland dataset at 10× magnification^13–17^.

As shown in **Table 3**, DermNet achieved the highest Dice score across all six segmentation classes: 0.98 for BKG, 0.95 for EPI, 0.92 for PAP, 0.94 for RET, 0.97 for KER, and 0.98 for HYP. Among the comparator models, TransUNet achieved the next-highest scores for EPI and PAP at 0.88 and 0.85, respectively, whereas SwinUNet achieved 0.87 and 0.84 and nnU-Net achieved 0.86 and 0.83 for these classes. These results demonstrate consistently higher segmentation performance for DermNet across both larger and more structurally challenging tissue compartments.

**Table 3.** Comparison of DermNet with state-of-the-art segmentation models on the Queensland dataset (10x magnification) for each segmentation class representing background (BKG), epidermis (EPI), papillary dermis (PAP), reticular dermis (RET), keratin (KER), and hypodermis (HYP).

| **Methodology** | **BKG** | **EPI** | **PAP** | **RET** | **KER** | **HYP** |
|---|---|---|---|---|---|---|
| UNet++ | 0.92 | 0.84 | 0.81 | 0.79 | 0.88 | 0.93 |
| DeepLabv3+ | 0.91 | 0.85 | 0.82 | 0.80 | 0.87 | 0.90 |
| TransUNet | 0.94 | 0.88 | 0.85 | 0.84 | 0.89 | 0.92 |
| SwinUNet | 0.95 | 0.87 | 0.84 | 0.82 | 0.88 | 0.91 |
| nnU-Net | 0.93 | 0.86 | 0.83 | 0.81 | 0.89 | 0.93 |
| **DermNet (Ours)** | **0.98** | **0.95** | **0.92** | **0.94** | **0.97** | **0.98** |

To determine whether the observed differences were statistically significant, pairwise Wilcoxon signed-rank tests were performed between DermNet and the competing segmentation models. As illustrated in **Figure 6**, all comparisons yielded *p* values below the predefined significance threshold of 0.05, indicating statistically significant differences in performance between DermNet and the evaluated comparator models.

**Figure 6.**
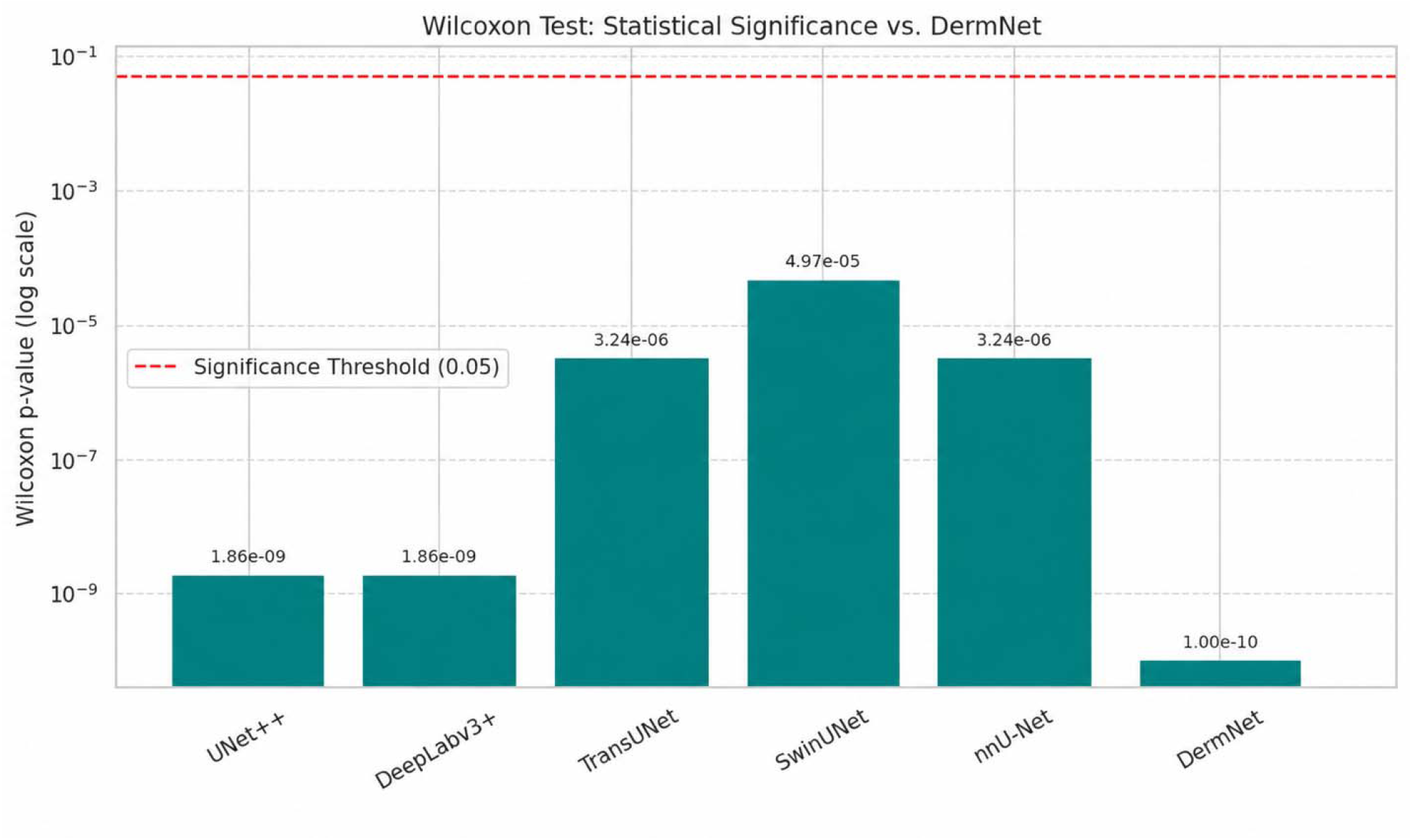
Statistical significance of performance differences between DermNet and competing segmentation models. Wilcoxon signed-rank test **p** values are shown on a logarithmic scale for comparisons with UNet++, DeepLabv3+, TransUNet, SwinUNet, and nnU-Net. The dashed horizontal line represents the statistical significance threshold (**p** = 0.05); values below this threshold indicate statistically significant differences.

The computational complexity and inference time of DermNet and the comparator architectures are presented in **Figure 7**, providing an additional comparison of model performance in relation to computational requirements.

**Figure 7.**
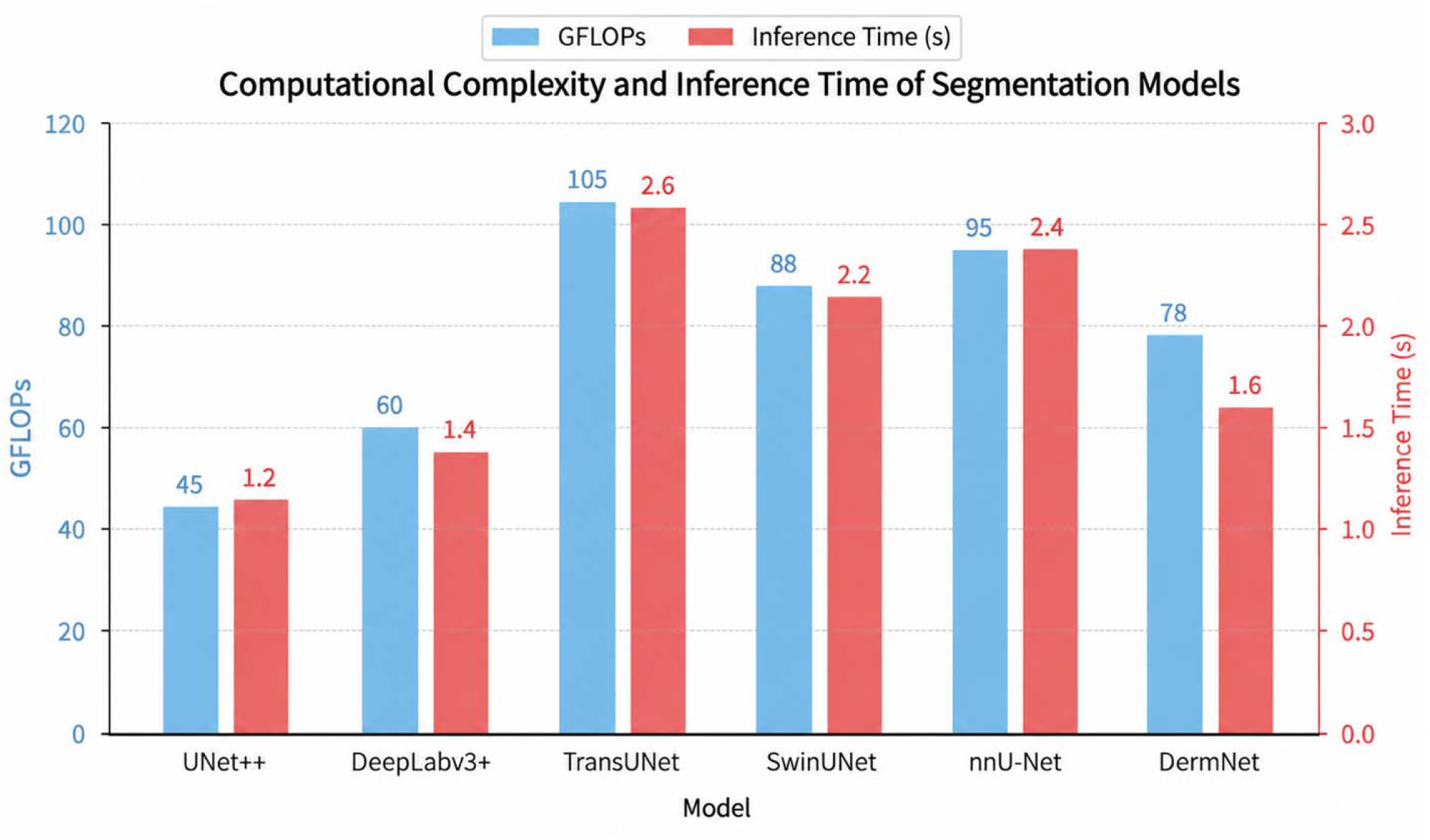
Computational complexity and inference time of DermNet compared with state-of-the-art segmentation models. Computational cost, measured in GFLOPs, and inference time (seconds) are compared across UNet++, DeepLabv3+, TransUNet, SwinUNet, nnU-Net, and DermNet, demonstrating the computational efficiency of DermNet relative to other segmentation architectures.

### Comparison with different loss functions

We conducted an empirical study by employing various combinations of the loss functions to evaluate the model’s performance. **Table 4** shows how well Model-III works with various sets of loss functions, and it is proof of how such combinations can increase the segmentation ability of skin tissue layers. From the results, it is found that the inclusion of complementary loss functions significantly boosts the model’s ability to learn complex patterns and, therefore, enhances the performance in terms of accuracy.

**Table 4.**
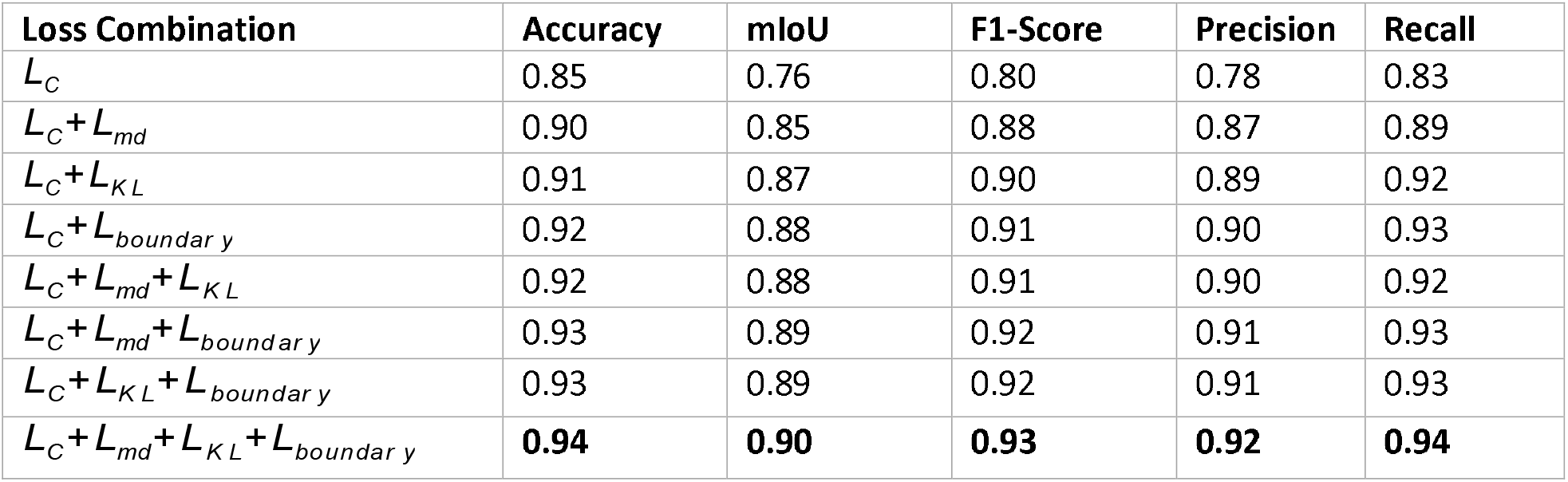
Performance evaluation of the proposed framework (Model-III) by combining various Loss Functions when trained and tested on the Entire Dataset (10x, 5x, 2x).

| Loss Combination | Accuracy | mIoU | F1-Score | Precision | Recall |
|---|---|---|---|---|---|
| *L*<sub>*C*</sub> | 0.85 | 0.76 | 0.80 | 0.78 | 0.83 |
| *L*<sub>*C*</sub>+*L*<sub>*md*</sub> | 0.90 | 0.85 | 0.88 | 0.87 | 0.89 |
| *L*<sub>*C*</sub>+*L*<sub>*KL*</sub> | 0.91 | 0.87 | 0.90 | 0.89 | 0.92 |
| *L*<sub>*C*</sub>+*L*<sub>*boundary*</sub> | 0.92 | 0.88 | 0.91 | 0.90 | 0.93 |
| *L*<sub>*C*</sub>+*L*<sub>*md*</sub>+*L*<sub>*KL*</sub> | 0.92 | 0.88 | 0.91 | 0.90 | 0.92 |
| *L*<sub>*C*</sub>+*L*<sub>*md*</sub>+*L*<sub>*boundary*</sub> | 0.93 | 0.89 | 0.92 | 0.91 | 0.93 |
| *L*<sub>*C*</sub>+*L*<sub>*KL*</sub>+*L*<sub>*boundary*</sub> | 0.93 | 0.89 | 0.92 | 0.91 | 0.93 |
| *L*<sub>*C*</sub>+*L*<sub>*md*</sub>+*L*<sub>*KL*</sub>+*L*<sub>*boundary*</sub> | **0.94** | **0.90** | **0.93** | **0.92** | **0.94** |

The classification loss (*L*_*C*_ ) alone already establishes a baseline performance with 0.85 accuracy and a mean Intersection over Union of 0.76. While (*L*_*C*_ ) works well for general classification, it does not tackle the intricacies associated with tissue structure segmentation, such as the keratin layer, for which more accurate boundary delineation is required. The mutual distillation loss *L*_*md*_ significantly improves the performance with an mIoU of 0.85 and a recall of 0.89. This loss function *L*_*md*_ promotes the sharing of knowledge among different branches of the model, thereby enhancing the learning of more robust and complete feature representations required for discrimination between subtle differences in tissue classes. Adding the Kullback-Leibler divergence loss, (*L*_*KL*_ ), fine-tunes the class probability distributions of the predictions, thus improving class discrimination and making more accurate predictions. The recall values achieve 0.92. The boundary loss ( *L*_*bound ar y*_ ) is crucial in the definition of fine details in tissue boundaries, especially for classes that have limited pixel representation, such as the keratin layer and epidermis. The addition of ¿ increases recall to 0.93 by reducing false negatives and allowing better segmentation of neighboring or overlapping classes.

By adding up ( *L*_*C*_ + *L*_*md*_+ *L*_*K L*_+ *L*_*bound ar y*_ ), the best performance is obtained at an accuracy of 0.94, mean IoU of 0.90, and F1-score of 0.93. Altogether, these loss functions allow the model to perceive general as well as specific characteristics of the skin tissue layers. These handle problems such as class imbalance, misalignment of probabilities, and ambiguity at boundaries.

### Comparison with Similar Incremental Learning Frameworks

We conducted further experiments about the performance evaluation of the proposed framework with other leading incrementally learning frameworks. Specifically, the comparison was performed between two frameworks: the first evaluation with the proposed model running with *L*_*wM*_ loss that replaces the general *L*_*K L*_ (KL divergence) loss with the *L*_*AD*_ (adversarial divergence) loss; and the second entry adds with the *L*_*EM*_ (embedding distillation) loss in addition to the losses of *L*_*CE*_ (cross-entropy) and *L*_*KD*_ (knowledge distillation).

**Table 5** shows the outcome efficiency of the models, indicating that the inclusion of *L*_*K L*_ makes the model able to converge better than other loss functions such as *L*_*AD*_ and *L*_*EM*_ in the alternative frameworks. The loss *L*_*K L*_ centers on the relative entropy or change in the probability distribution of the extracted features and enables the model to concentrate on the importance of each feature in the new learning process. This results in a stronger learning trajectory for this model as retaining important features while discarding irrelevant ones is essentially encouraged. As evident from the results of Model-III, which adopted *L*_*K L*_, yielded maximum accuracy (95.55%), mIoU (93.72%), and F1-score (95.42%) are obtained using all loss function combinations; thus, indicating *L*_*K L*_ is one of the contributing factors to the model’s superior performance.

**Tabel 5.** Comparative performance of different loss functions across the incremental learning stages. Model-I, Model-II (FL), and Model-III represent the three sequential stages of incremental training. Performance is reported in terms of accuracy, mean Intersection over Union (mIoU), F1-score, forward transfer (FWT), backward transfer (BWT), and forgetting score (FS). MICIL and \(L_{wM}\) represent the alternative loss-based methodologies evaluated against the proposed DermNet framework. Bold values indicate the best-performing results within each comparison. FL, feature learning; FS, forgetting score; FWT, forward transfer; BWT, backward transfer; mIoU, mean Intersection over Union.

| **Model** | **Methodology** | **Accuracy** | **mIOU** | **F1-Score** | **FWT** | **BWT** | **FS** |
|---|---|---|---|---|---|---|---|
| Model-I |  | 0.88 | 0.87 | 0.89 | 0 | 0 | 0 |
| Model-II(FL) | **MICIL** | 0.91 | **0.90** | 0.88 | 0.01 | -0.01 | 0.5 |
| Model-III |  | 0.95 | 0.93 | 0.95 | -0.05 | -0.05 | 0.5 |
| Model-I |  | 0.88 | 0.87 | 0.89 | 0 | 0 | 0 |
| Model-II(FL) | *L*<sub>*wM*</sub> | 0.90 | 0.89 | **0.90** | 0.002 | -0.03 | 0.4 |
| Model-III |  | 0.92 | 0.91 | 0.91 | -0.08 | -0.04 | 0.3 |
| Model-I |  | 0.88 | 0.87 | 0.89 | 0 | 0 | 0 |
| Model-II(FL) | **DermNet** | **0.93** | 0.90 | **0.90** | 0.5 | 0.01 | 0.0001 |
| Model-III |  | **0.97** | **0.94** | **0.96** | 0.8 | 0.02 | 0 |

As evident from **Table 5**, *L*_*AD*_ loss in *L*_*wM*_ framework does provide good improvements, however, does not achieve the highest result. This is because *L*_*AD*_ focuses on a very simple minimization of the adversarial divergence while the real task is rather about relative entropy, where the actual measure of difference comes in the definition between two feature distributions, thus making it relatively less accurate in guiding the model through the learning.

Additionally, the *L*_*EM*_ loss when profused with Model-III according to its workability fails to provide the same amount of improvement as does *L*_*K L*_ for the present study. Although Model-II (FL), with *L*_*EM*_ as the underlying loss function, gives stellar results in mIoU (90.50), performance metrics for Model-III using *L*_*K L*_ leave all others behind, producing both higher mIoU and F1 score (94.83 and 96.74, respectively). Ultimately, results in Table 5 speak volumes about the importance of the proposed framework utilizing *L*_*K L*_ loss in combination with the rest of the loss functions to achieve better performance than when including either *L*_*AD*_ or *L*_*wM*_ .

### Evaluating DermNet on other Publicly Available Dataset

To evaluate generalizability beyond the development dataset, DermNet was tested on the independent Histo-Seg: H&E Whole Slide Image Segmentation Dataset, comprising 38 H&E-stained WSIs with corresponding histopathologist-generated ground-truth annotations.

As presented in **Table 6**, DermNet maintained high segmentation performance on the independent dataset, with class-wise Dice scores ranging from 0.89 for PAP to 0.97 for BKG. EPI and RET, which represent relatively thin and morphologically complex tissue regions, achieved Dice scores of 0.92 and 0.93, respectively. Corresponding IoU and recall values were also consistently high across the evaluated classes.

**Table 6.** Performance of DermNet on another publicly available dataset for each segmentation class representing background (BKG), epidermis (EPI), papillary dermis (PAP), reticular dermis (RET), keratin (KER), and hypodermis (HYP).

| **Class** | **Dice** | **IoU** | **Recall** | **BD-Score** |
|---|---|---|---|---|
| BKG | 0.97 | 0.96 | 0.98 | 0.96 |
| EPI | 0.92 | 0.90 | 0.93 | 0.91 |
| PAP | 0.89 | 0.90 | 0.92 | 0.90 |
| RET | 0.93 | 0.92 | 0.91 | 0.91 |
| KER | 0.95 | 0.93 | 0.91 | 0.94 |
| HYP | 0.90 | 0.92 | 0.93 | 0.92 |

These results demonstrate that the segmentation performance observed on the development dataset was largely maintained when DermNet was evaluated on an independent histopathological dataset.

### Qualitative Evaluation and Model Interpretability

To evaluate the model qualitatively, we have inferred the model using sample images from test split comprising all magnification levels (10x, 5x, 2x). Additionally, we also computed heat maps and t-SNE plots to visualize the model’s insight and to add explainability to the conducted research. Additionally, medical imaging requires high precision and recall, therefore we plot the receiver operating characteristic (ROC) curve to emphasize high precision and recall which adds high value to the proposed framework.

### Multi-Resolution Segmentation

Figure 8. presents representative qualitative segmentation results across the three magnification levels. Panel (a) shows the input H&E-stained WSIs, panel (b) the corresponding ground-truth annotations, and panels (c), (d), and (e) DermNet predictions at 2×, 5×, and 10×, respectively.

Across magnifications, the predicted segmentation masks showed close visual correspondence with the ground-truth annotations. Predictions at 10× magnification demonstrated finer delineation of tissue structures and boundaries, consistent with the greater availability of fine-grained histological detail. At lower magnifications, some loss of boundary sharpness and minor misclassification were observed; however, the overall morphology and spatial organization of the principal tissue compartments remained preserved.

**Figure 8.**
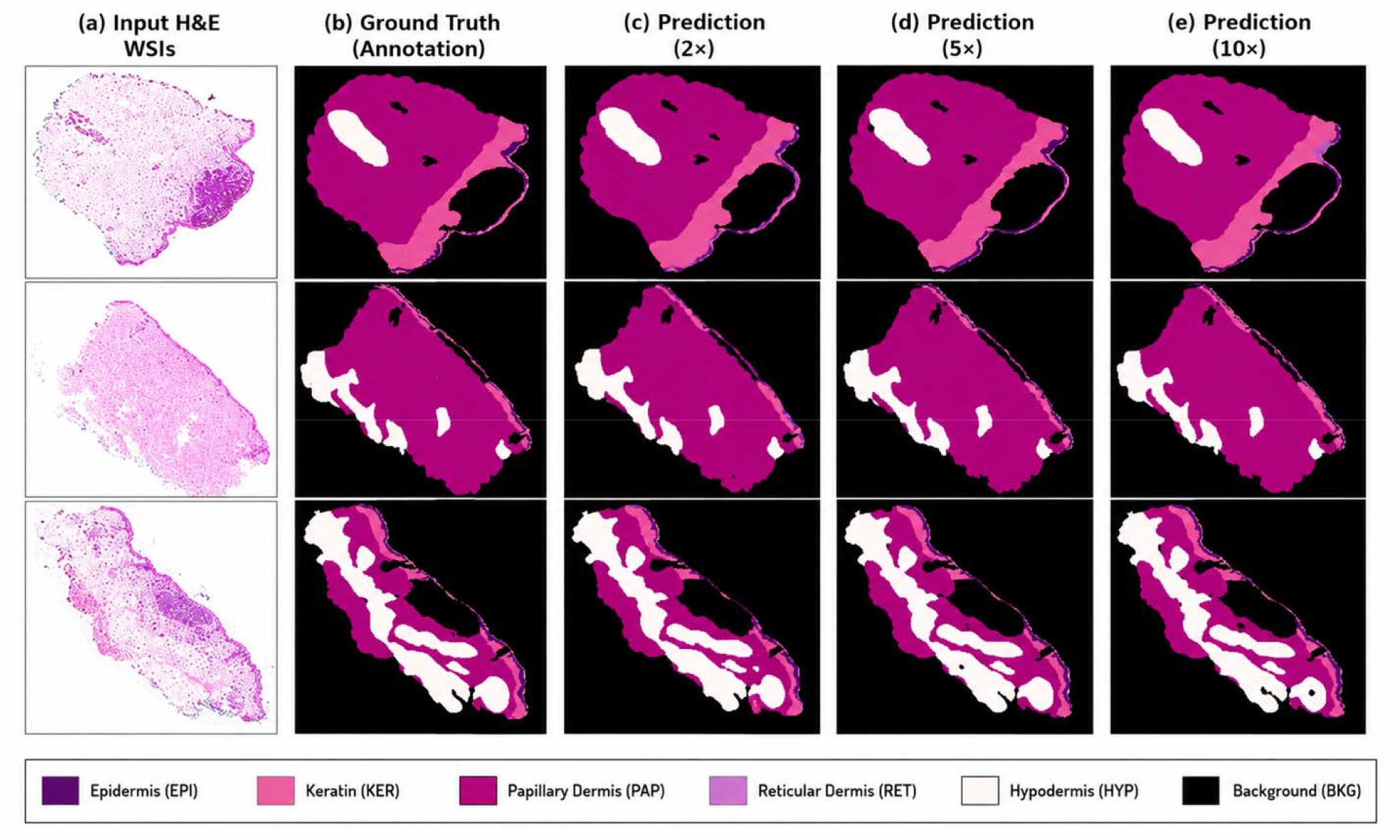
Qualitative evaluation of DermNet for skin layer segmentation across multiple magnification levels. (a) Representative H&E-stained whole-slide images, (b) corresponding ground-truth annotations, and DermNet segmentation outputs at (c) 2×, (d) 5×, and (e) 10× magnifications. The predictions demonstrate preservation of overall tissue morphology across resolutions, with finer boundary delineation at higher magnification.

These qualitative findings complement the quantitative multi-resolution results and demonstrate the ability of a single DermNet framework to generate segmentation outputs across the three evaluated magnification levels.

### Class-Wise Activation Maps

Class-specific activation maps were generated from the fully trained DermNet model to examine the image regions contributing to individual tissue-class predictions. As shown in **Figure 9**, segmentation masks and corresponding activation heatmaps were generated for EPI, HYP, KER, RET, PAP, and BKG. Warmer regions of the activation maps represented areas of greater model attention, whereas cooler regions represented lower relevance to the corresponding class prediction. Across the evaluated classes, regions of stronger activation generally corresponded spatially to the relevant segmented tissue compartments. These maps provide a qualitative assessment of the spatial features contributing to DermNet’s class-specific predictions.

**Figure 9.**
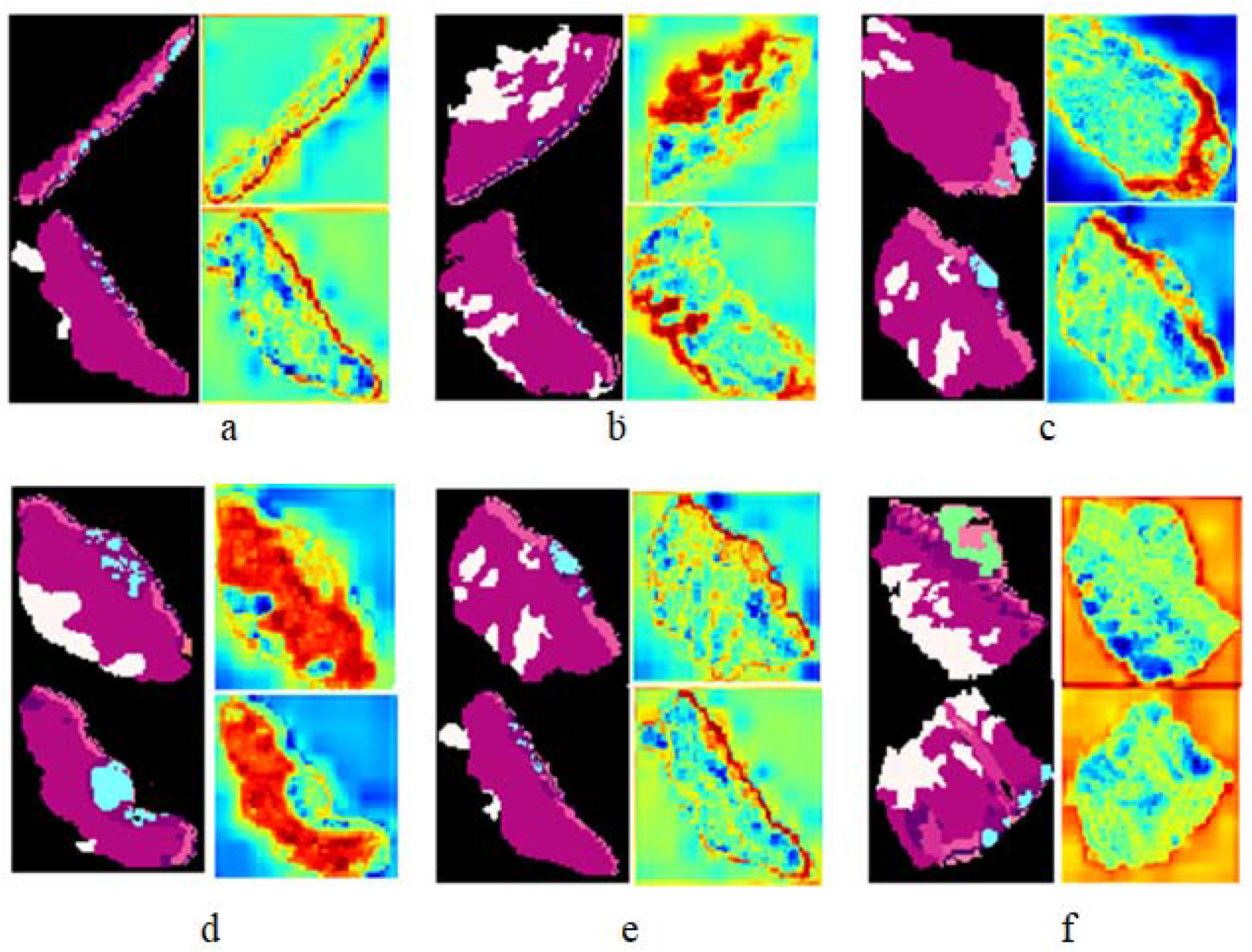
Class-wise activation maps generated by DermNet for skin layer segmentation. Segmentation masks and corresponding activation heatmaps demonstrate the regions contributing to model predictions for (a) epidermis, (b) hypodermis, (c) keratin, (d) reticular dermis, (e) papillary dermis, and (f) background. Warm regions indicate greater model attention, whereas cooler regions indicate lower relevance to the corresponding class prediction.

### Precision–Recall and ROC Analysis

Class-wise precision–recall and ROC analyses were performed for BKG, HYP, RET, PAP, EPI, and KER. As illustrated in **Figure 10**, DermNet maintained high precision over a broad range of recall values across the six tissue classes. The class-wise curves showed generally consistent performance, including for morphologically complex classes such as KER and HYP.

**Figure 10.**
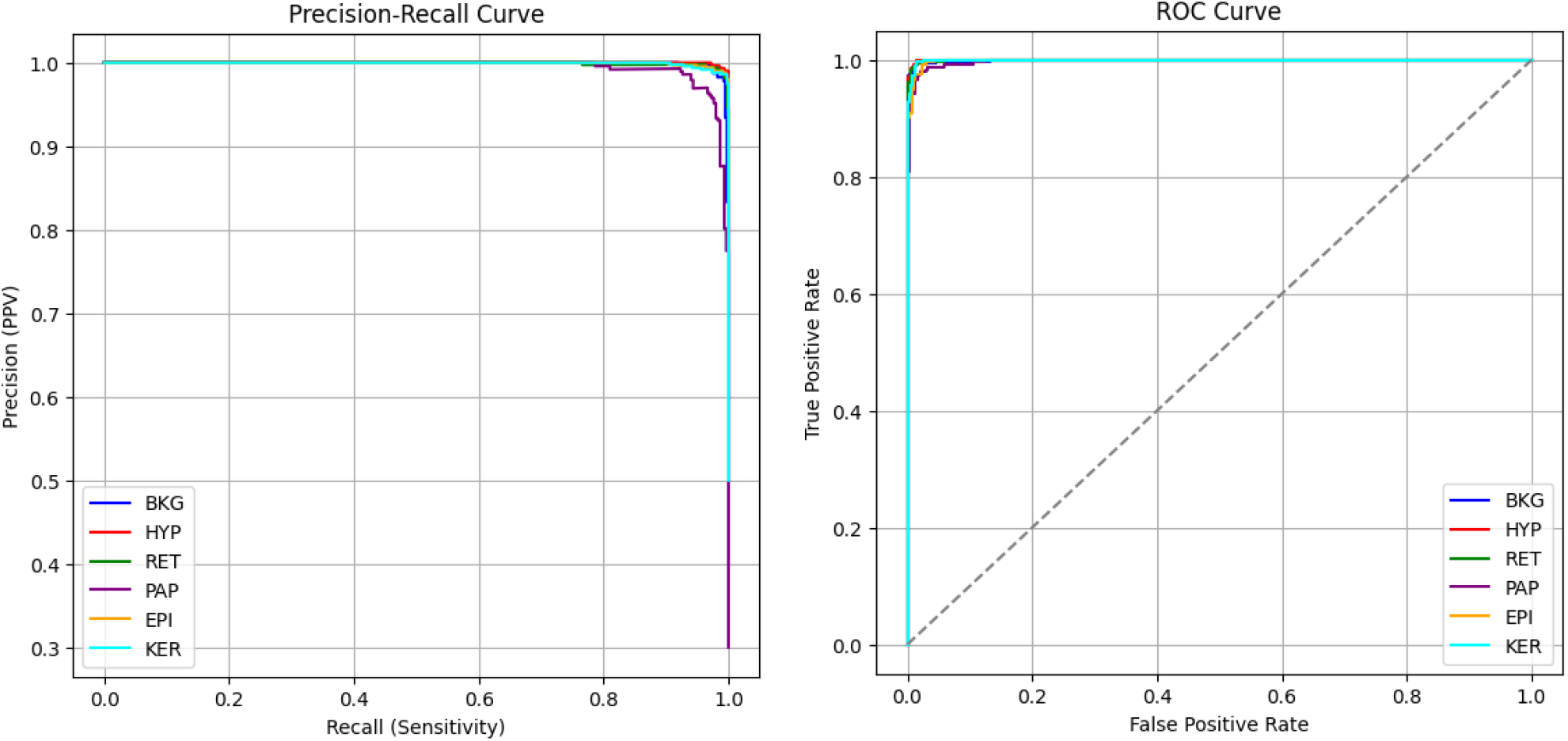
Class-wise precision–recall and receiver operating characteristic (ROC) curves for DermNet. Curves are shown for background (BKG), hypodermis (HYP), reticular dermis (RET), papillary dermis (PAP), epidermis (EPI), and keratin (KER), demonstrating high precision, recall, sensitivity, and specificity across the segmented classes.

A decline in precision was observed toward the highest recall values for some classes, indicating a limited number of more ambiguous predictions at extreme operating thresholds. Overall, the precision– recall and ROC analyses were consistent with the class-wise segmentation metrics and demonstrated strong discrimination across the evaluated tissue compartments.

### Boundary Accuracy

To further evaluate segmentation boundary accuracy, we computed HD for each tissue class, as illustrated in **Figure 11**. The boxplots show the distribution of HD values for KER, HYP, PAP, BKG, RET, and EPI, where lower values indicate greater agreement between the predicted and ground-truth boundaries.

**Figure 11.**
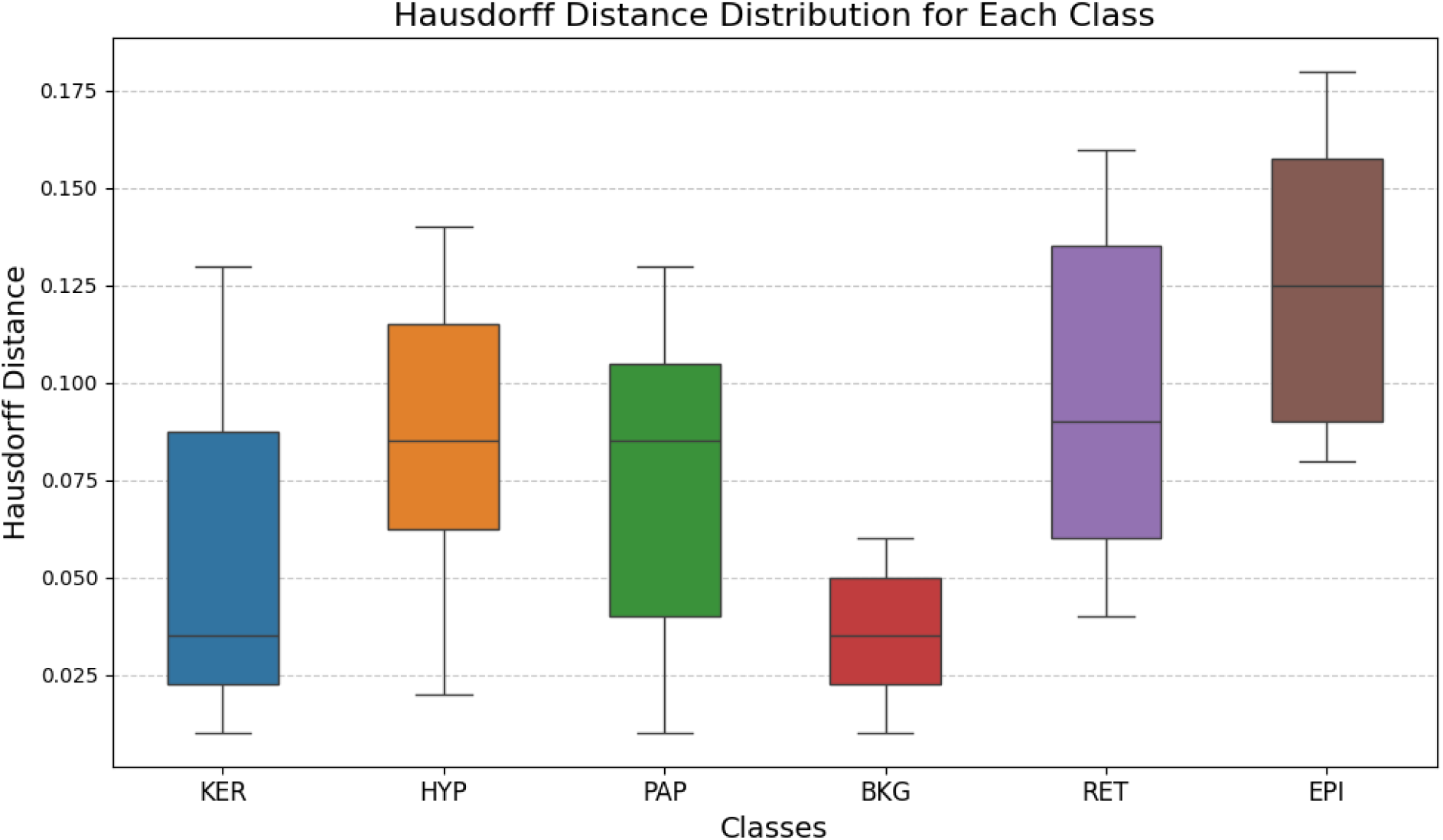
Class-wise distribution of Hausdorff Distance for DermNet segmentation. Boxplots illustrate the distribution of Hausdorff Distance values for keratin (KER), hypodermis (HYP), papillary dermis (PAP), background (BKG), reticular dermis (RET), and epidermis (EPI). Lower Hausdorff Distance values indicate closer agreement between predicted and ground-truth segmentation boundaries.

### Feature Visualization and Model Discriminability

The learned feature space was visualized using t-SNE to examine the distribution of representations corresponding to the six segmentation classes. As shown in **Figure 12**, features formed class-associated clusters with relatively limited intermixing between several tissue categories.

**Figure 12.**
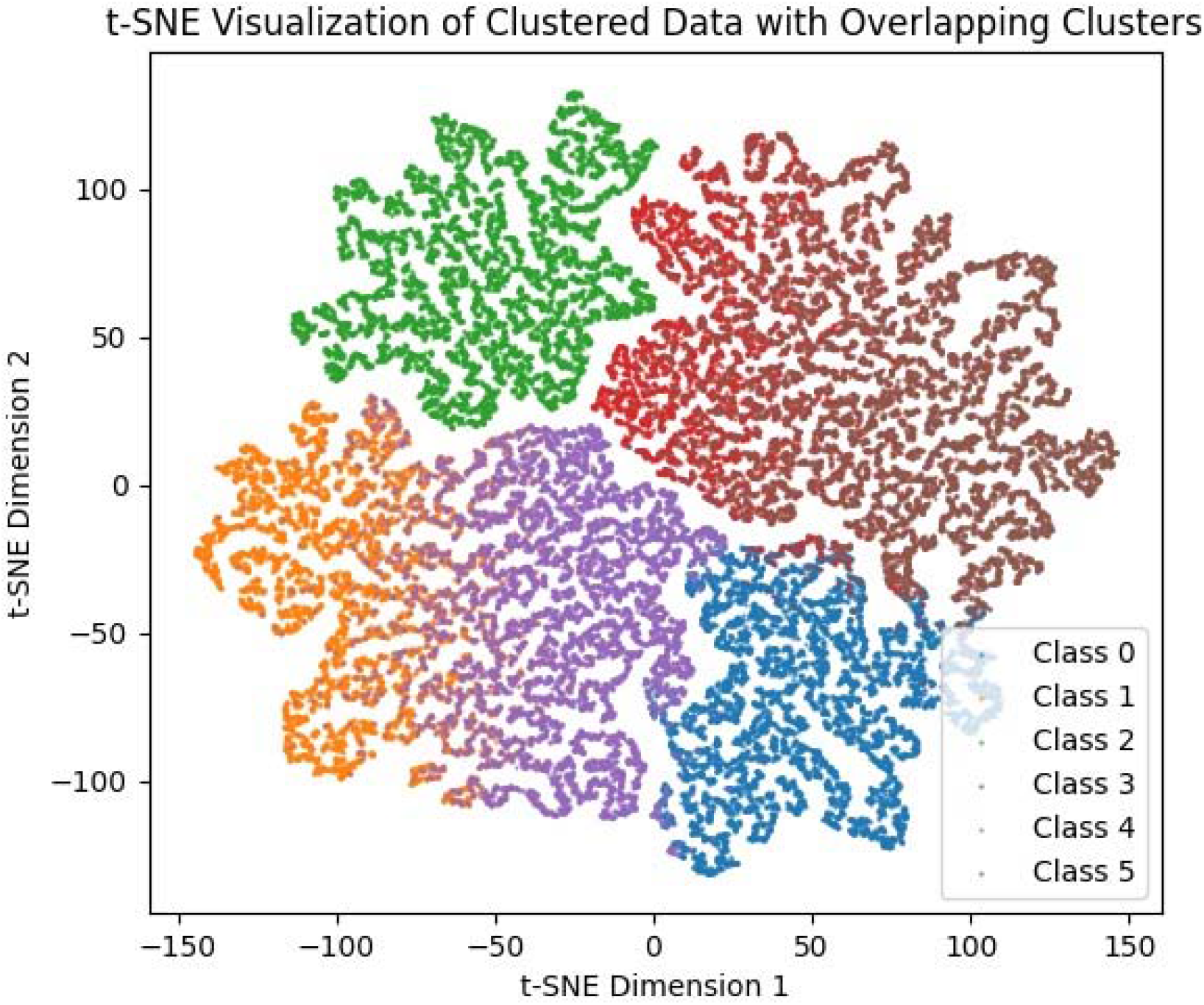
t-SNE visualization of feature representations learned by DermNet. The two-dimensional projection illustrates the clustering of learned features across the six segmentation classes: background (BKG; Class 0), epidermis (EPI; Class 1), papillary dermis (PAP; Class 2), reticular dermis (RET; Class 3), hypodermis (HYP; Class 4), and keratin (KER; Class 5). The distribution demonstrates class-specific feature clustering with varying degrees of overlap between morphologically related tissue classes.

BKG (Class 0), EPI (Class 1), PAP (Class 2), RET (Class 3), HYP (Class 4), and KER (Class 5) demonstrated varying degrees of separation within the two-dimensional feature space. Compact clustering was observed for several classes, whereas partial overlap remained between morphologically related tissue compartments. Overall, the visualization indicates that DermNet learned distinguishable feature representations for the evaluated skin tissue classes.

## Discussion

In this study, we developed DermNet, a multi-resolution class-incremental learning framework for automated segmentation of skin layers in histopathological WSIs. DermNet progressively learns epidermis, papillary dermis, reticular dermis, keratin, and hypodermis while incorporating images acquired at 10×, 5×, and 2× magnifications. The proposed framework demonstrated consistently strong segmentation performance across tissue classes, outperformed conventional and state-of-the-art segmentation architectures, and maintained high performance on an independent public dataset. These findings support the utility of combining multi-resolution representation learning with incremental knowledge transfer for complex dermatopathological segmentation.

Accurate delineation of skin structures is clinically relevant because the localization and distribution of pathological features across the epidermis, dermis, and deeper tissues contribute to histopathological diagnosis. Previous studies have demonstrated that segmentation of clinically meaningful structures in skin biopsy WSIs can facilitate automated tissue analysis and downstream diagnostic classification^7,18,19^. The publicly available dataset used in the present study provides densely annotated WSIs encompassing multiple skin compartments and non-melanoma skin cancers, creating an important resource for developing automated dermatopathology algorithms^8^. Unlike approaches primarily designed to identify tumors or individual structures, DermNet was developed to distinguish multiple anatomically related skin layers and tissue compartments within a unified segmentation framework.

A key feature of DermNet is its incorporation of multiple magnification levels. Histopathological interpretation inherently requires assessment of both global tissue architecture and fine morphological detail. Multi-resolution computational pathology approaches have similarly demonstrated that combining contextual information obtained at lower magnification with fine-grained information at higher magnification can improve semantic segmentation. For example, HookNet integrates information from different resolutions and has demonstrated advantages over single-resolution approaches in histopathological tissue segmentation^20^. More recent work has further supported multi-resolution deep learning for extracting biologically meaningful structures from H&E-stained WSIs^21^. DermNet extends this concept to skin layer segmentation by incorporating 10×, 5×, and 2× WSIs throughout incremental training, allowing the model to learn complementary tissue representations across spatial scales. Another important contribution is the application of class-incremental learning to skin layer segmentation. Conventional deep learning models are generally trained simultaneously on all available classes and may require substantial retraining when new classes or datasets are introduced. Continual learning instead enables models to acquire information sequentially, although catastrophic forgetting of previously learned representations remains a central challenge^22,23^. DermNet addresses this problem through exemplar replay, knowledge distillation, and EWC, enabling previously learned class representations to be retained while new tissue classes are progressively introduced. The favorable FWT, BWT, and FS observed with DermNet further support the effectiveness of this strategy in balancing acquisition of new information with retention of previously learned representations.

The ablation experiments further demonstrated the contribution of the proposed optimization strategy. Classification loss alone established baseline segmentation performance, whereas sequential incorporation of mutual distillation, KL divergence, and boundary loss improved the evaluated performance metrics. These components provide complementary objectives: mutual distillation facilitates knowledge transfer between successive models, KL divergence promotes consistency between probability distributions, and boundary loss emphasizes spatial agreement along tissue interfaces. Boundary-aware optimization is particularly relevant in skin histology, where thin or morphologically adjacent structures may have complex and irregular interfaces. The superior performance of the complete loss formulation therefore suggests that combining class-level knowledge transfer with boundary-sensitive optimization contributes to accurate skin layer segmentation.

The findings are further supported by the model’s external validation. DermNet maintained strong segmentation performance on the independent Histo-Seg dataset, with class-wise Dice scores ranging from 0.89 to 0.97, suggesting that the learned representations were not restricted to the development dataset. This is important because differences in tissue preparation, staining, scanning, and institutional workflows can introduce domain variability in computational pathology. Continual-learning research increasingly recognizes such distributional changes as an important challenge for the clinical translation of medical imaging models^22–24^. Although evaluation on a single independent dataset does not establish broad clinical generalizability, the preservation of performance across datasets provides preliminary evidence of the robustness of DermNet to variations in histopathological images.

The explainability analyses provide additional insight into model behavior. Class-specific activation maps demonstrated spatial correspondence between areas of greater model attention and the relevant tissue compartments, while ROC and precision–recall analyses demonstrated strong class discrimination. The t-SNE visualization further demonstrated class-associated clustering of learned representations across the six segmentation classes. Although these analyses cannot independently establish clinical interpretability or reliability, they provide complementary evidence that DermNet learns class-specific histological representations rather than relying exclusively on nonspecific image characteristics.

Beyond segmentation performance, DermNet may have potential applications within digital dermatopathology workflows. Automated delineation of tissue compartments could facilitate downstream quantitative analysis, disease localization, and computer-assisted diagnosis while potentially reducing the time required for manual tissue annotation. The use of multiple magnification levels also more closely reflects the multiscale assessment performed during conventional histopathological examination. However, the present findings should be considered a demonstration of technical performance rather than evidence of improved clinical workflow or diagnostic accuracy; these potential benefits require prospective evaluation.

Despite these promising findings, several limitations should be acknowledged. First, DermNet was developed using a relatively limited public dataset of 290 WSIs, although the dataset provides detailed annotations across multiple tissue types and biopsy formats^8^. Second, class imbalance remains inherent to histopathological segmentation because tissue compartments occupy substantially different proportions of WSIs. Third, although external validation supports generalizability, broader evaluation across independent institutions, scanners, staining protocols, biopsy types, and disease populations is necessary before clinical implementation. Fourth, the present study evaluates segmentation performance rather than improvement in prospective diagnostic decision-making or pathologist efficiency. Last, the exemplar-replay strategy requires retention of previously encountered samples, which may introduce storage and data-governance considerations on larger clinical datasets.

Future studies should therefore evaluate DermNet using larger, multicenter datasets and investigate strategies for addressing class imbalance and domain variability. Continual WSI analysis has demonstrated the feasibility of sequentially adapting models to new datasets and tasks, supporting further investigation of this approach in computational pathology^23–25^. Incorporating additional histopathological classes and imaging modalities may further improve the framework’s applicability and generalizability. Prospective studies should also determine whether automated skin layer segmentation can improve downstream disease classification, quantitative biomarker assessment, diagnostic accuracy, or pathologist efficiency.

## Conclusion and Future Work

The proposed DermNet framework demonstrates that class-incremental learning can enable accurate and robust segmentation of skin layers from whole slide images (WSIs). By combining mutual knowledge distillation with multi-resolution learning at 10×, 5×, and 2× magnifications, DermNet progressively learns class-specific features while preserving previously acquired knowledge and mitigating catastrophic forgetting. The combination of complementary loss functions further enhances feature representation and segmentation performance, while the ablation studies, class-wise performance metrics, activation maps, and t-SNE analysis support the robustness and reliability of the framework. DermNet also has potential applications in digital dermatopathology by facilitating automated tissue characterization, reducing diagnostic workload, and supporting more consistent analysis of WSIs. Its multi-resolution approach reflects the evaluation of tissue at different magnifications routinely performed by pathologists and may therefore facilitate integration into digital pathology workflows. Future studies should focus on addressing class imbalance, validating the framework across larger and more diverse multicenter datasets, and incorporating additional imaging modalities to further improve its generalizability and clinical applicability.

## Abbreviations

ADE20K: Ade20k dataset
BCC: Basal cell carcinoma
BD-Dice: Boundary dice
BKG: Background
BWT: Backward transfer
CAD: Computer-aided diagnosis
CNN: Convolutional neural network
EPI: Epidermis
EWC: Elastic weight consolidation
FIM: Fisher information matrix
FOL: Follicle
FS: Forgetting score
FWT: Forward transfer
H&E: Hematoxylin and eosin
HYP: Hypodermis
IEC: Intraepidermal carcinoma
INF: Inflammation
IoU: Intersection over Union
KER: Keratin
KL: Kullback–Leibler
mIoU: Mean Intersection over Union
PAP: Papillary dermis
RET: Reticular dermis
ROC: Receiver operating characteristic
SCC: Squamous cell carcinoma
t-SNE: T-Distributed Stochastic Neighbor Embedding
WSI: Whole slide image

## Appendix

## Acknowledgments

The authors have no additional acknowledgments to declare.

## Author contributions statement

Salam AA: Conceptualization of this study, Methodology, Software, Writing-Original draft. Akram MU: Formal Analysis, Supervision, Writing-Review & Editing. Jawed S: Methodology, Software, Writing-Original draft. Shehryar T: Methodology, Software, Writing-Original draft. Fazal ZZ: Writing-Original draft, Writing-Review & Editing. Yousaf MH: Validation, Writing-Review & Editing. Rao B: Investigation, Data Curation, Writing-Review & Editing. Alghamdi NS: Supervision, Software, Writing-Original draft. All authors reviewed and approved the final manuscript.

## Data Availability

Data utilized in this research is a public repository “Histopathology Non-Melanoma Skin Cancer Segmentation Dataset” available at https://doi.org/10.14264/8be4bd0.

## Ethics Approval and Informed Consent

This study involved secondary analysis of data obtained from publicly available, de-identified datasets and did not involve recruitment, intervention, or direct interaction with human participants. Therefore, institutional review board approval and informed consent were not required for the present study.

